# Towards an extended classification of noise-distortion preferences by modeling longitudinal dynamics of listening choices

**DOI:** 10.1101/2024.10.25.24316092

**Authors:** Giulia Angonese, Mareike Buhl, Jonathan A. Gößwein, Birger Kollmeier, Andrea Hildebrandt

## Abstract

Individuals have different preferences for setting hearing aid (HA) algorithms that reduce ambient noise but introduce signal distortions. “Noise haters” prefer greater noise reduction, even at the expense of signal quality. “Distortion haters” accept higher noise levels to avoid signal distortion. These preferences were assumed to be stable over time, and individuals were classified solely on the basis of these stable, trait scores. However, the question remains as to how stable individual listening preferences are and whether day-to-day state-related variability needs to be considered as a further criterion for classification. We designed a mobile task to measure noise-distortion preferences over two weeks in an ecological momentary assessment study with *N* = 185 (106 f, *M_age_* = 63.1, *SD_age_* = 6.5) unaided individuals with subjective hearing difficulties. Latent State-Trait Autoregressive (LST-AR) modeling was used to evaluate stability and dynamics of individual listening preferences. The analysis revealed a significant amount of state-related variance. The model has been extended to a mixture LST-AR model for data-driven classification, taking into account trait and state components of listening preferences. In addition to successful identification of noise haters, distortion haters and a third intermediate class based on longitudinal, outside of the lab data, we further differentiated individuals with different degrees of variability in listening preferences. It follows that individualisation of HA fitting could be improved by assessing individual preferences along the noise-distortion trade-off, and the day-to-day variability of these preferences needs to be taken into account for some individuals more than others.

## Introduction

Proper fitting of hearing aids (HAs) is critical to user satisfaction, speech intelligibility, and listening comfort. Noisy situations are among the most challenging environments for HA users (Perry et al., 2019). Therefore, noise reduction (NR) algorithms are an essential part of the HA, as they aim to improve listening comfort and speech intelligibility by improving the signal-to-noise ratio (SNR) in noisy situations and reducing the annoyance of ambient noise. However, the increase in listening comfort comes at the expense of speech naturalness. NR processing introduces a certain amount of distortion into the speech signal due to noise estimation errors and processing artifacts of the algorithm (Brons, Dreschler, et al., 2014; Houben et al., 2012; Reinten et al., 2019). Such signal degradation reduces speech intelligibility and perceived sound quality (Arehart et al., 2007).

Individual preferences for NR settings are subject to large between-person variability (Brons et al., 2013; Brons, Houben, et al., 2014; Houben et al., 2023; Kubiak et al., 2022; Völker et al., 2018). Some individuals prefer weak NR (i.e., accept lower listening comfort) to minimize signal distortions and have been referred to as “NR haters” (Neher & Wagener, 2016) or “distortion haters” (Kubiak et al., 2022; Völker et al., 2018). Conversely, “noise haters” (Kubiak et al., 2022; Völker et al., 2018) also called “NR lovers” (Neher & Wagener, 2016), prefer aggressive NR despite the presence of more distortions that impact speech naturalness. An additional intermediate third category has also been found in recent studies (Houben et al., 2023; Kubiak et al., 2022), including individuals that prefer a more moderate NR strength.

The underlying determinants of these individual preferences are still being investigated. While some studies suggest that individuals with larger pure tone average (PTA) prefer stronger NR (noise haters) (Houben et al., 2023; Neher & Wagener, 2016), this relationship has not been confirmed in other studies (Brons, Dreschler, et al., 2014; Kubiak et al., 2022). Instead, the Speech Recognition Threshold (SRT) turned out to better predict individual listening preferences than other individual factors like PTA (Kubiak et al., 2022). Effects of HA use on NR preferences have not yet been investigated, though first findings point towards more tolerance of signal distortions for experienced HA users, potentially due to adaptation to HA signal processing (Reinten et al., 2023). Self-report questionnaires (sound preference and hearing habits) or cognitive performance measures (reading span) did not explain between-person variability in NR preferences (Neher, 2014; Neher et al., 2016).

At the within-person level, listening preferences for different HA settings (e.g., intensity, gain-frequency slope and directionality) have been shown to vary across different auditory environments and situational contexts (Korzepa et al., 2018; Walravens et al., 2020). Under similar listening conditions, individual listening preferences have so far been considered as prototypical subjective traits, stable over time with good retest reliability (Nelson et al., 2018; Völker et al., 2018). With respect to NR algorithms, experienced HA users were found to exhibit consistent preferences for NR strength at a test-retest over a one year period (Neher & Wagener, 2016). Stability of individual preferences along the noise-distortion trade-off was also observed in a one-week test-retest (Kubiak et al., 2022), as well as over three (Reinten et al., 2023) and six (Houben et al., 2012) consecutive repetitions for both normal-hearing and hearing-impaired individuals. However, to our knowledge, no study investigated the stability of listening preferences across repeated measures on multiple consecutive days.

According to the revised Latent State-Trait (LST) theory (Steyer et al., 2015), observations do not occur in a situational vacuum. This requires to account not only for measurement errors, but also for situational (or state) fluctuations of the personal attribute (trait) to be measured. Longitudinal data modelled within the LST framework can provide important insights into the degree of stability (i.e., consistency of the trait) and within-person variability (i.e., state-related variance) of individual listening preferences. Indeed, a recent study which demonstrated reliable day-to-day fluctuations of hearing performance (Kuhlmann et al., 2023) calls for the assessment of potential fluctuations in individual listening preferences for noise vs. distortion, given their association with auditory measures.

The development of a mobile measure of NR strength preference would facilitate repeated assessment of listening preferences outside the lab and in different listening situations, thereby increasing knowledge on their potential within-person variability. Moreover, such a mobile task would pave the way for future implementations of self-adjustment options for NR algorithms in hearing mHealth applications. The observed inter-individual differences in listening preferences underscore the need of self-tailored designs and personalized HA solutions (Nielsen et al., 2018).

In the present study, we developed an objective and mobile measure of individual listening preferences along the noise-distortion trade-off, based on artificially introduced, controlled distortions. To assess trait-as well as state-related characteristics, we collected longitudinal data on listening preferences as part of a larger Ecological Momentary Assessment (EMA) study. The Latent State-Trait Autoregressive (LST-AR) model (Holtmann et al., 2023) was used to quantify the temporal dynamics of individual listening preferences. This model has been further extended to a mixture LST-AR model to identify latent subgroups of individuals that differ in their listening preferences, both related to trait and state properties. In addition, we tested for associations of listening preferences with different trait-related covariates (hearing performance, self-reported sound preferences, noise sensitivity and personality traits).

More specifically, the following research aims and questions were addressed:

RQ1. Are the psychometric properties of the proposed individual listening preferences measure for noise vs. distortion applied on a smartphone acceptable?
RQ2. Evaluate trait consistency and state specificity of listening preferences: is the trait stable or does it fluctuate considerably over time (state-related variance)?
RQ3. Perform a data-driven, model-based classification of noise and distortion haters:

3.1 Can individuals be classified based on their trait levels measured by the mobile app?
3.2 Can we further differentiate the classification when taking the latent state variability into account?
3.3 Which covariates are associated with these latent classes? The purpose of this analysis is to gain a better understanding of the class properties.

## Methods

### Study overview

The data was collected in the context of an EMA study distributed over three consecutive weeks, as described in Angonese et al. (2024). The study was conducted in the field using the personal smartphones of the participants. During the first week, participants were asked to complete different questionnaires on their smartphone browser once a day. This baseline assessment aimed at collecting information on several audiological and psychological variables associated with hearing and hearing help-seeking (Knoetze et al., 2023). In the present study, we considered a subset of the baseline questionnaires, namely sound preferences and hearing habits, noise sensitivity and personality. For further measures included, please refer to Angonese et al. (2024). During the second and third week, participants completed a longitudinal assessment of listening preferences (noise-distortion trade-off task) and hearing performance in their everyday life. A total of 20 measurement time points were distributed along 10 weekdays (morning and evening) and lasted approximately 15 minutes each. The study was conducted using formr, an open-source web-based application programming interface for the `R` language that creates automated studies (Arslan et al., 2019). Participants were prompted to conduct the experimental tasks via email and SMS and were remunerated with 10 Euros per hour. Written informed consent was collected prior to enrolment. The study plan and data management have been approved by the Research Ethics Committee of the Carl von Ossietzky Universität Oldenburg (08.09.2021, EK/2020/020-01).

### Participants

The study included *N* = 185 participants, 106 females and 79 males (0 diverse), aged between 47 and 82 years, with *M_age_* = 63.1 and *SD_age_* = 6.5. To target potential future users of HAs and hearing mHealth applications, we recruited older adults with self-reported hearing difficulties but without HAs. As part of the inclusion criteria, all participants were German native speakers and could use a smartphone. Less than half of the participants (48.1%) had previously consulted a doctor about their hearing problems, which had been present for an average of 5.2 years. One third of the sample reported the presence of tinnitus (35.1%).

## Materials

### Mobile assessment of listening preferences

We designed a mobile task in formr to assess listening preferences along the trade-off between noise annoyance and signal distortion, simulating the general effect of a NR algorithm. The stimuli and the task were chosen for comparability with previous studies (Gößwein et al., 2022; Kubiak et al., 2022). The task is based on sliders with discrete buttons, along which the SNR (and artificially introduced distortions based on peak clipping) between a speech and a noise signal can be varied. By comparing the results of different sliders, the trade-off between noise and distortion preferences will be assessed as explained in the following sections. After one practice trial, the task was presented three consecutive times at baseline to obtain a multiple indicator trait measure. The task was presented an additional 20 times during the longitudinal phase of the study.

### Stimuli

The stimuli consisted of speech signals in background noise, with or without distortions contingent on the task condition. The signals were constructed as follows.

- **Speech signal.** The target speech material consisted of sentences from the German Matrix Sentence Test (Oldenburger Satztest – OLSA, Kollmeier et al., 2015) with female, synthetic speech (Nuesse et al., 2019). The sentences were pseudo-randomised to differ across stimuli. The level of the speech signal was varied during the experiment, indicated in the following by the SNR relative to the noise signal.
- **Noise signal.** The noise consisted of multi-talker babble noise (Kubiak et al., 2022), and its level was kept fixed across stimuli. This noise is a mix of 10 adult voices (male and female) superimposed to the point of unintelligibility, with a total duration of 60 seconds. For each stimulus, a different random noise snippet corresponding to the respective duration of the sentence was extracted from the noise file.
- **Clipping.** Signal distortions were introduced by applying hard peak clipping to the speech signal similar to Kubiak et al., 2022. The clipping rate was defined as the number of clipped speech samples divided by the total number of speech samples. The Perceptual Similarity Measure (PSM) of the PEMO-Q model (Huber & Kollmeier, 2006) was used to achieve perceptually equidistant distortions over stimuli, such that the clipping can be varied approximately linearly with the SNR (along the slider dimension, see below). The PSM measure predicts the difference in perceived audio quality between the reference signal (the unclipped mixed signal, with noise and speech) and the test signal (the clipped mixed signal, with noise and clipped speech). To achieve a linear perception of distortions with increasing SNR (in the respective SNR range for each slider), the target quality (PSM) was estimated by a linear fit to PSM values obtained for linear clipping values (in the interval [0 to 80] %) employed as starting point. After that, the optimal clipping values (supplementary material 1) were estimated by interpolation to the respective target PSM, based on precalculated PSM maps for all combinations of SNR and clipping values. In addition to the sound quality, we modelled the perceived loudness by means of the loudness model of Moore and Glasberg (ISO 532-2). This was necessary in order to equalize the loudness of all stimuli since in general clipping increases the loudness of a signal. The loudness of the unclipped mixed signal was used as reference loudness.

### Noise-distortion trade-off task

The noise-distortion trade-off task included three listening conditions, each presented in the form of a slider with nine discrete positions. A speech-in-noise stimulus of approximately two seconds (length of the respective sentence) was presented at each slider position. The three listening conditions were adapted from measures previously used in laboratory settings (Gößwein et al., 2022; Kubiak et al., 2022) and are summarized in figure 1.

1. **Simple linear gain condition (slider 1).** This condition identified the SNR level at which the individual reports little listening effort in the absence of signal distortions. Nine signals were presented with linear gain applied to the speech signal, namely with increasing SNR from left to right (in the interval [−7 to 23] dB SNR in steps of 3.75 dB), with varying speech level and constant noise level as adjusted to a comfortable level by subjective calibration (see below under *Procedure*). The range of SNR values provided a complete range of listening effort. The lower limit was chosen to avoid too high listening effort, as values below −7 dB SNR would require excessive listening effort. Individuals with normal hearing associated an SNR between −7 and 22 dB with a “moderate listening effort”, with 22 dB SNR being mostly “effortless” (Krueger et al., 2017).
2. **General trade-off condition (slider 2).** In this condition, better SNR comes at the expense of hard peak-clipping distortions. As in slider 1, the nine stimuli had linearly increasing SNR from left to right (in the interval [−7 to 23] dB SNR in steps of 3.75 dB). In addition, hard peak clipping of the signal was introduced as perceptually linearly increasing from left to right. Thus, increasing SNR corresponded to increasing clipping distortions. The peak clipping rate ranged from 0 to 80% of the samples clipped, in 10% steps according to the button spacing.
3. **Adaptive trade-off condition (slider 3).** This last condition was similar to slider 2, but here the SNR range was dependent on the participant’s SNR value selected in slider 1. The minimum value for the SNR remained −7 dB SNR (leftmost position) and it linearly increased up to 3 dB above the SNR value measured with slider 1. As an exception, if the participants selected the initial two values on slider 1 (−7 or −3.25 dB SNR) the upper-limit of slider 3 was set at 3 dB SNR. If they selected the last value on slider 1 (23 dB SNR), the upper limit of slider 3 was kept at 23 dB SNR. Clipping distortions of the signal were introduced as in slider 2 and increased perceptually linear from left to right (in the interval [0 to 80] %), thus with a different mapping of clipping values to SNR. This adaptive SNR range has been implemented in order to enforce the trade-off between noise and distortion by providing a higher amount of distortion at a SNR level that was previously judged to require little listening effort (slider 1).

**Figure 1.**
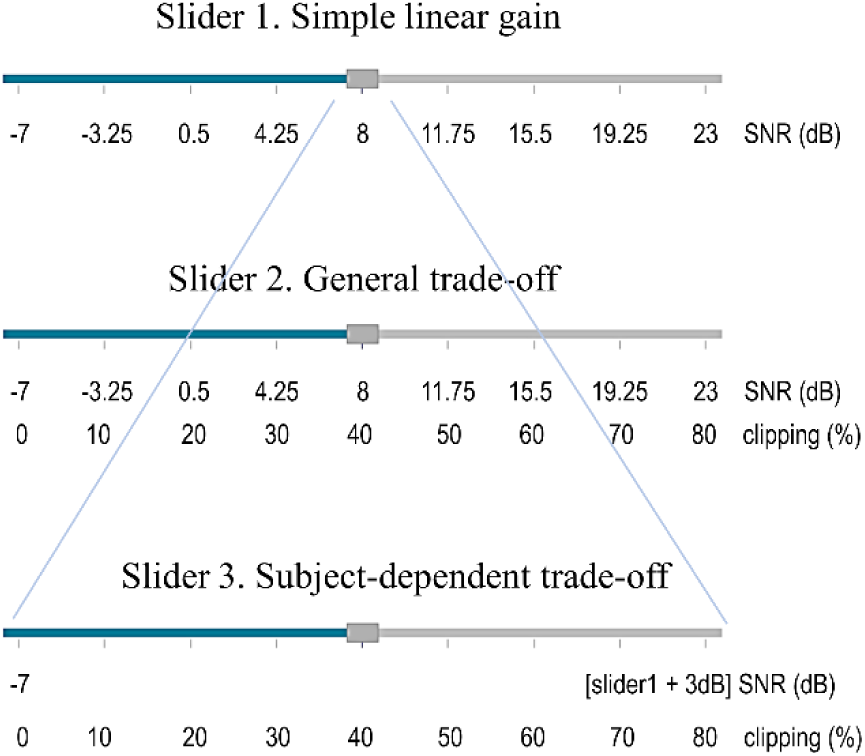
Graphical representation of the three listening conditions, which were presented to participants in the form of sliders. Each slider consisted of nine discrete positions, each containing a speech-in-noise stimulus. Information on SNR and clipping values for each position on the slider is shown. SNR: signal-to-noise ratio.

### Procedure

Participants were instructed to perform the task on their smartphone browser in a quiet environment. They were asked to use their personal headphones and to refrain from changing them throughout the study. It should be noted that we did not enquire about the type/brand of smartphone and headphones used. Three participants used the smartphone loudspeakers, as headphones were not available to them. A pilot study was conducted in June 2021 with 21 participants (10 males, 11 females, aged between 18 and 70 years old) to develop appropriate and clear task instructions. At each task presentation (before slider 1), users were asked to perform a subjective calibration by adjusting the noise stimulus to a comfortable listening level. This level was defined as the smartphone’s output volume with headphones (due to implementation limitations of formr we could not control this level via the software). Participants were instructed to keep this volume level constant during the entire task. Users were asked to move the slider button stepwise from left to right via the touchscreen until they could hear the speech with little listening effort. Figure 2 shows how the task was displayed on the smartphone’s browser (see also supplementary material 2).

**Figure 2.**
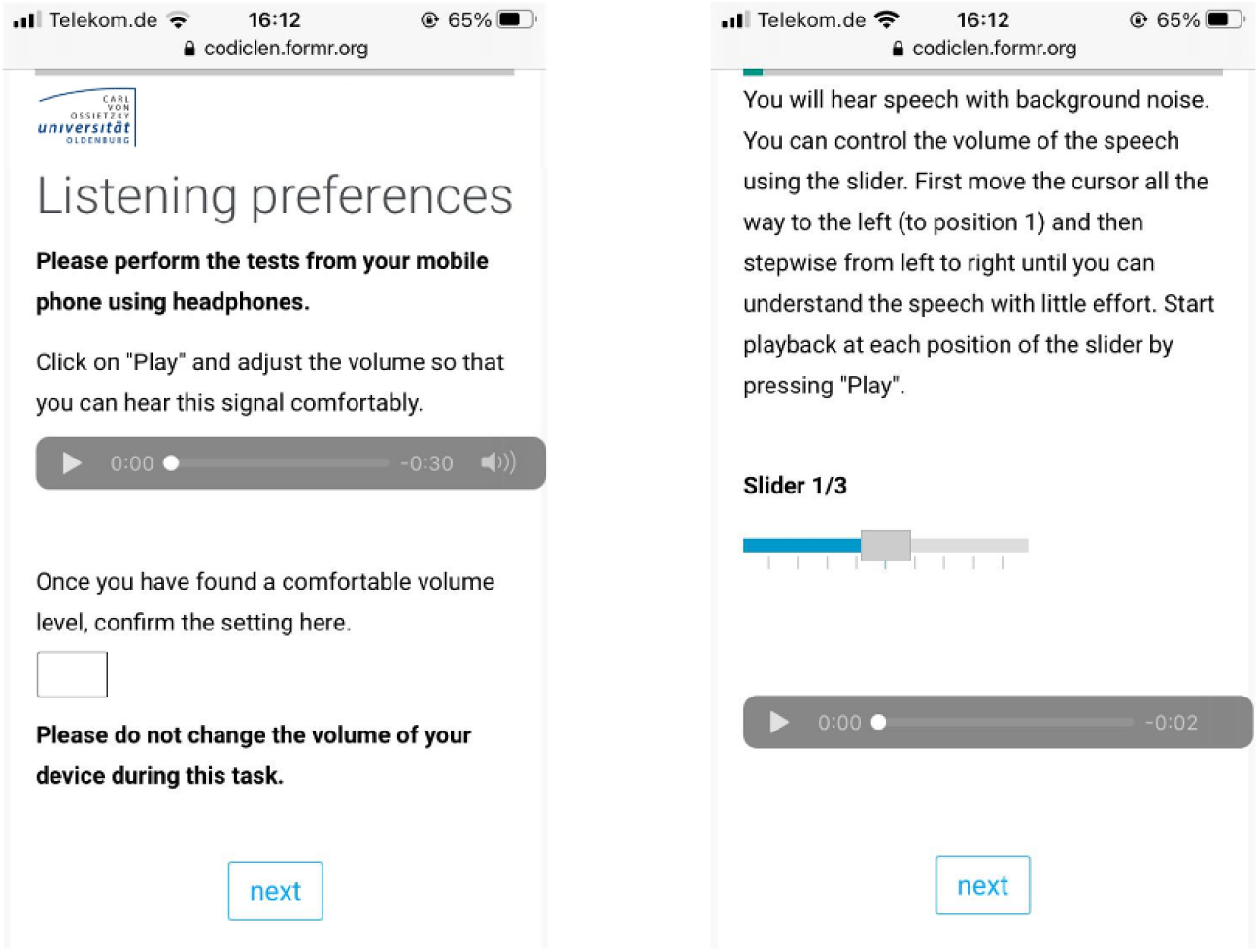
Task design as displayed on the participants’ smartphone. First, participants were asked to complete a calibration task to adjust the volume level of their device (left). The three sliders followed, each in a separate browser page. Here, only slider 1 is shown (right). Supplementary material 2 provides an overview of the entire task (all three sliders) as displayed on the browser. The instructions have been here translated to English for consistency with the language of the paper, but were presented to the study participants in German.

### Measures of listening preferences for noise vs. distortions

Two measures were derived from the noise-distortion trade-off task: The SNR difference between the response to slider 1 and 2, namely between the simple linear gain vs. the general trade-off condition (ΔSNR_g_), and the SNR difference between slider 1 and 3, namely between the simple linear gain vs. adaptive trade-off condition (ΔSNR_a_). Both measures aimed to assess the preferred trade-off between noise and distortions at the individual point of lowest listening effort. The goal of the psychometric analysis (RQ1) was to investigate which of the two difference measures was more consistent at the between-person level and better able to capture the targeted trade-off, that is, providing suitable combinations of SNR and clipping that enforce a decision between the two dimensions. Figure 3 provides an example of ideal preferences along each slider for a noise hater and a distortion hater. Noise haters would show a ΔSNR_g,a_ close to zero, meaning that they desire a similarly high SNR in slider 2/3 and slider 1, at the expense of a high level of distortions causing a degradation in speech quality. In an ideal scenario, a noise hater would choose the same SNR level on all sliders (note that we looked at the differences between slider 1 and 2 as well as slider 1 and 3). On the contrary, distortion haters would show a positive ΔSNR_g,a_, indicating that they accept a lower SNR in slider 2/3 as compared to slider 1 in order to avoid distortions. This choice would ensure better speech quality, albeit at the expense of listening effort and speech intelligibility performance at lower SNR. As illustrated in figure 3, the constrained SNR range of slider 3 forces a distortion hater to select an even lower SNR compared to slider 2, given that the clipping value at the same button position is higher for slider 3. Accordingly, we anticipate that the trade-off would be more evident in ΔSNR_a_ measures. It is important to note that even distortion haters would prefer a high SNR if distortions were to be removed. Indeed, depending on the degree of hearing loss, a distortion hater can sacrifice some SNR to avoid distortions only at the point where speech intelligibility is still acceptable.

**Figure 3:**
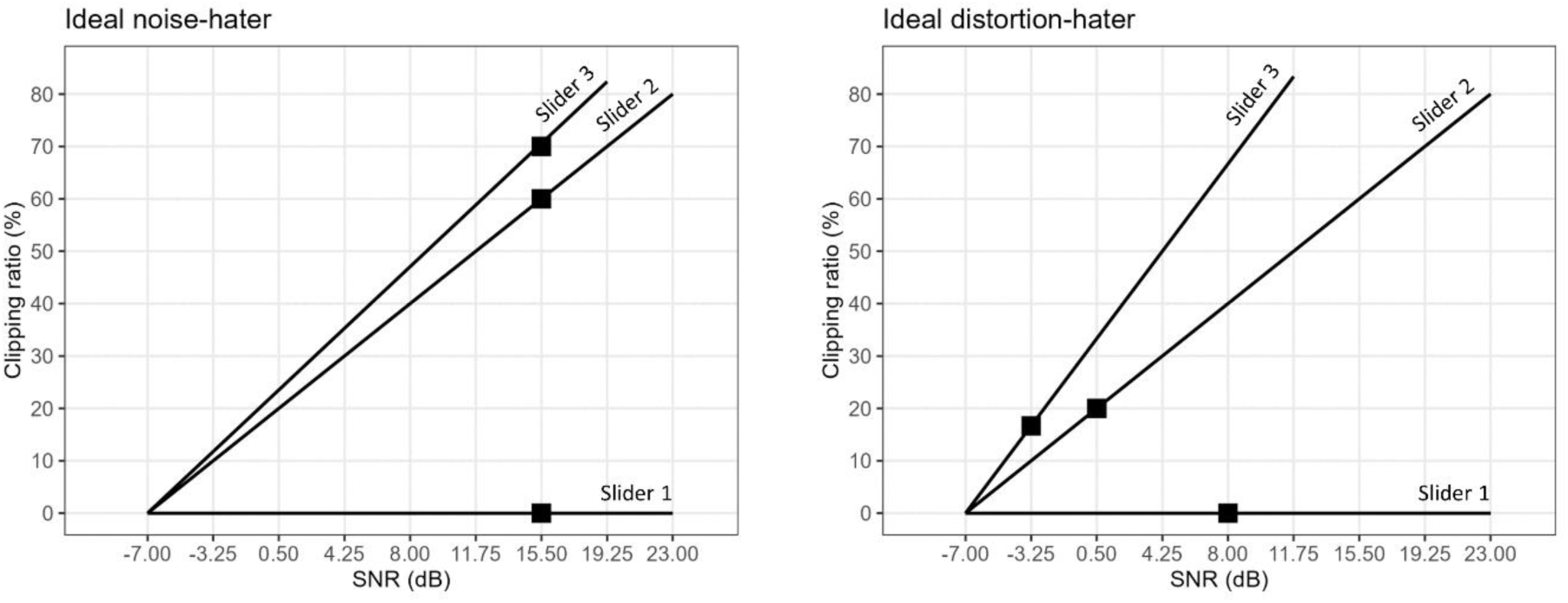
Graphical representation of ideal slider position choices (black squares) for a noise hater (left) and a distortion hater (right). The two graphs show the SNR values (X axis) and clipping values (Y axis) for slider 1 (simple linear gain condition), slider 2 (general trade-off condition) and slider 3 (adaptive trade-off condition). Here it can be seen how slider 3 values vary depending on the choice made in slider 1, while slider 1 and 2 are the same for all participants.

### Assessment of trait-related covariates

#### Hearing performance

Hearing performance was assessed with the Digit Triplets Test (DTT) (Smits et al., 2004; Buschermöhle et al., 2014), an adaptive speech-in-noise screening test that measures SRT with triplets of monosyllabic digits presented in test-specific noise (i.e., superimposed from speech material). The DTT has proven to be suitable for mobile, remote self-test screening of hearing abilities due to its robustness to ambient noise levels outside of audiometric booth environments (Denys et al., 2019). Moreover, SRT estimation is relatively robust against changes in presentation level and no exact calibration is needed (Löw et al., 2018). Thanks to its low linguistic and cognitive demands the DTT is also a valid instrument for the older population (Denys et al., 2019). The test was performed in an online browser page hosted by the Hörzentrum Oldenburg gGmbH. Participants were instructed to use their personal headphones and to keep the same type of headphones throughout the study. Three participants used phones-loudspeakers due to reported technical difficulties. The use of different types or quality of headphones has shown no impact on test reliability for the DTT (Potgieter et al., 2016; Van den Borre et al., 2021). Each hearing test began with a signal adjustment trial, in which a digit triplet in noise was presented to the participants with the task to “adjust the volume to hear both digits and noise clearly”. After a practice trial, participants completed 20 daily hearing assessments during the longitudinal part of the study. The hearing performance data was then summarized into an individual trait score (mean SRT).

#### Sound preference and hearing habits

The German version of the Sound Preference and Hearing Habits Questionnaire (SPHHQ) (Meis et al., 2018) was used at baseline to assess the individual sound preferences profile, which can be seen as a stable trait related to preferences for specific sounds and hearing habits. Its 23 items load onto seven different factors. Three of them have been included here as potential covariates, as they have been considered as mostly related to response to noise and processing artifacts (Neher et al., 2016): annoyance or distraction by background noise; importance of sound quality; noise sensitivity. Higher scores on each scale reflect higher agreement with the respective domain.

#### Noise sensitivity

Noise sensitivity is a stable, individual attribute that influences reactions to environmental sounds and indicates how sensitive a person is to perceived noise. Individual sensitivity to perceived noise was assessed through the Weinstein Noise Sensitivity Scale (WNSS) (Weinstein, 1978; Zimmer & Ellermeier, 1997). It consists of 21 items investigating reactions and attitudes towards noise in general and in relation to environmental sounds. The global score was used as covariate, with a higher global score indicating greater noise sensitivity.

#### Personality

The German version of the NEO Five-Factor Inventory (NEO-FFI) (Borkenau & Ostendorf, 1993) was used at baseline to assess individual differences in the five major personality traits. From these, neuroticism, extraversion and conscientiousness have been considered as potential covariates. Neuroticism is the predisposition to experience negative emotions (Cox et al., 2005). Extraversion refers to the tendency to be optimistic and outgoing. Conscientiousness is associated with being proactive, organized and methodical (Cox et al., 2005). People high in neuroticism would be expected to show greater variability in their daily listening preferences, whereas people high in conscientiousness would be expected to show less variability. Recent research also suggests that higher levels of neuroticism and lower levels of extraversion explain higher listening effort and lower acceptable noise levels (Wöstmann et al., 2021).

### Statistical analysis

#### Data preprocessing

Data analysis was performed with `R` version 4.2.1 (R Core Team, 2019) and `Mplus` version 8.6 (B. O. Muthén & Muthén, 1998). Raw data from the questionnaires and the listening preference task was imported from the online platform formr into the `R` environment using the package formr (Arslan, 2018). The daily hearing test results were received via automatized emails from the Hörzentrum Oldenburg and imported in `R` as .eml files. In some cases, participants performed the hearing test more than once at a given measurement time point due to limitations of the formr implementation. When this happened, only the last SRT result at a given time point was kept for analysis. A total of 43.8% of participants completed all 20 hearing tests, and in 95.1% of cases at least 15 SRT results were obtained. By visual inspection of the individual SRT distributions, some specific outlier patterns were identified and removed (implausible SRT value at the first measurement time point). We reached 100% completeness rate for the baseline data.

Two data sets were used in the subsequent analysis. One data set included the baseline measures for all *N_1_* = 185 participants. These included the SPHHQ, WNSS and NEO-FFI questionnaires, together with the individual mean SRT and the three baseline repetitions of the noise-distortion task. A second data set contained the longitudinal measures of listening preferences (20 repetitions). Here, a visual inspection of slider responses in the noise-distortion trade-off task revealed careless or inattentive responding (Curran, 2016) in 16 participants, who showed no variance in their data. These participants were therefore excluded from the longitudinal data analysis, resulting in *N_2_* = 169 participants. In this second data set, 2.8% of data was NA.

#### RQ 1. Are the psychometric properties of the proposed individual listening preferences measure for noise vs. distortion applied on a smartphone acceptable?

For this analysis we used the baseline data (three consecutive repetitions of the task) with *N_1_* = 185 participants. The initial step was to assess the consistency of measurements for the two slider measures ΔSNR_g_ and ΔSNR_a_, which we will refer to as indicators. This analysis aimed at testing whether the two indicators measure the same rank order of individuals and, consequently, whether they would be equally suitable for assessing the noise-distortion trade-off. We run a confirmatory factor analysis (CFA) by fitting a model series with the `cfa`() function of the lavaan package in `R`. Two model versions were evaluated (figure 4). A two-factor model was initially fitted, with two separate latent factors *T*_Δg_ for ΔSNR_g_ and *T*_Δa_ for ΔSNR_a_. This model assumes the two indicators to measure at least partly different latent variables. Subsequently, a single-factor solution was evaluated by fixing the correlation between the two factors to one. This model assumes that both indicators measure the exact same latent variable *T*_Δ_. The `anova`() function applied to the model results was used to compute a χ^2^-difference test to compare the two models with regard to their fit. The following factor model equation (see also figure 4) applies:

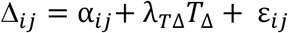

where Δ_*ij*_ is the measurement observation of indicator *i* (ΔSNR_g_ or ΔSNR_a_) at repetition *j* (for a total of 3 repetitions at baseline for each indicator); α_*ij*_ indicates the intercept and ε_*ij*_ the measurement error for each observation; *λ*_*T*Δ_ is the loading for the latent trait factor *T* of the variable ΔSNR. In a second step, after identifying the best-fitting model, we evaluated the factor loadings for ΔSNR_g_ and ΔSNR_a_ to identify which of the two slider measures should be considered for subsequent analysis, due to higher consistency. Higher loadings *λ*_*T*Δ_ of one measure on the latent variable indicate higher consistency of the measure between person.

**Figure 4.**
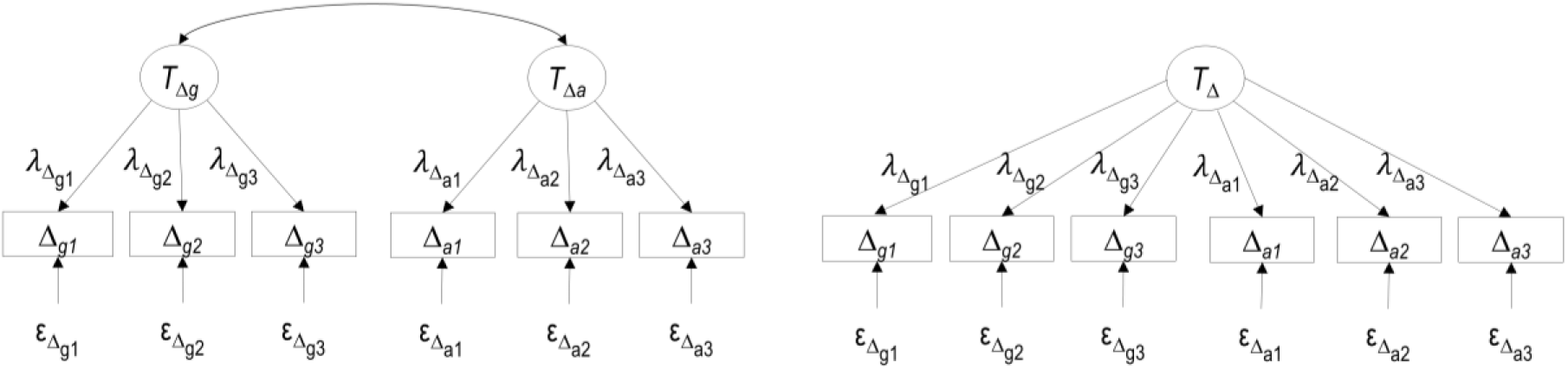
CFA diagrams of the model series for the two indicators ΔSNR_g_ (here depicted as Δ_g_) and ΔSNR_a_ (here depicted as Δ_a_) for the three measurement occasions at baseline (*N_1_ = 185*). (Left) Two-factor model. (Right) Single-factor model. *T*: latent trait; *λ*: factor loading; *Δ*: observed indicator; ε: measurement error.

#### RQ 2. Are listening preferences stable or do they fluctuate considerably over time (state-related variance)?

For the second research question, the longitudinal data frame (*N_2_* = 169 participants) was used, with up to 20 observations per participant. Temporal dynamics of individual listening preferences were investigated with a simplified version of the LST-AR model from Holtmann et al. (2023). In this modeling framework, individual short-term variability of latent processes can be modelled while taking into account temporal dependencies between consecutive observations (autoregressive effects). According to the LST theory, an observed variable *Δ_t_* (at time point *t*) can be decomposed into a trait variable *T_t_* (the attribute of a person (Steyer et al., 2015)), a residual variable *S_t_* (the attribute of a person in a situation (Steyer et al., 2015)), and a measurement error variable *ε_t_*. Trait and state variables together constitute a person’s true score. The trait is defined as the expectation of the person-specific distribution of *Δ_t_* across all possible situations that might be experienced at that time point (Holtmann et al., 2023). The state residual captures the effects of the situation and of the interaction between person and situation at a specific time point.

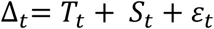

By extending the LST model to the LST-AR model, we consider carryover effects across temporally adjacent observations. The autoregressive (AR) parameters capture the effect of cumulative experiences across time, namely to which degree person-situation interaction effects at a time point *t* predict trait values at the following time point (Holtmann et al., 2023).

In our model, depicted in figure 5, we have only one indicator per time point (either ΔSNR_g_ or ΔSNR_a_ following RQ1). Hence, the residual variance which is not captured by the trait will include both the state-related variance (*S_t_*) and the error term (*ε_t_*). Further parameters introduced in the model equation are the intercept *α_t_*, which captures the average additive trait change, and the factor loadings **λ*_Tt_*, which have been constrained here to 1 under the assumption of time invariance. Our final model equation for each observation Δ_*t*_ is:

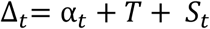

In a first step, a single-trait LST model (without AR effects) has been implemented for the indicator of choice (based on the results of RQ1) in `Mplus` using the Maximum likelihood robust (MLR) estimator with 100 random sets of starting values. The trait mean together with intercepts and residual variances were estimated for each time point. We evaluated trait consistency and state specificity by estimating the amount of state-related variance. A large proportion of state-related variance would indicate that listening preferences are not stable over time. In a second step, the model has been extended to a LST-AR model to gain more insight on potential time-series dynamics. In this step, we evaluated the temporal linear dependence of an observation on the previous time point, defined as first-order AR effect.

Model fit was evaluated using the root-mean-square error of approximation (RMSEA), standardised root mean square residual (SRMR), comparative fit index (CFI), Tucker-Lewis index (TLI), and the χ^2^-test, together with Akaike information criterion (AIC), Bayesian information criterion (BIC) and sample-size adjusted BIC (SABIC) for model comparison.

**Figure 5.**
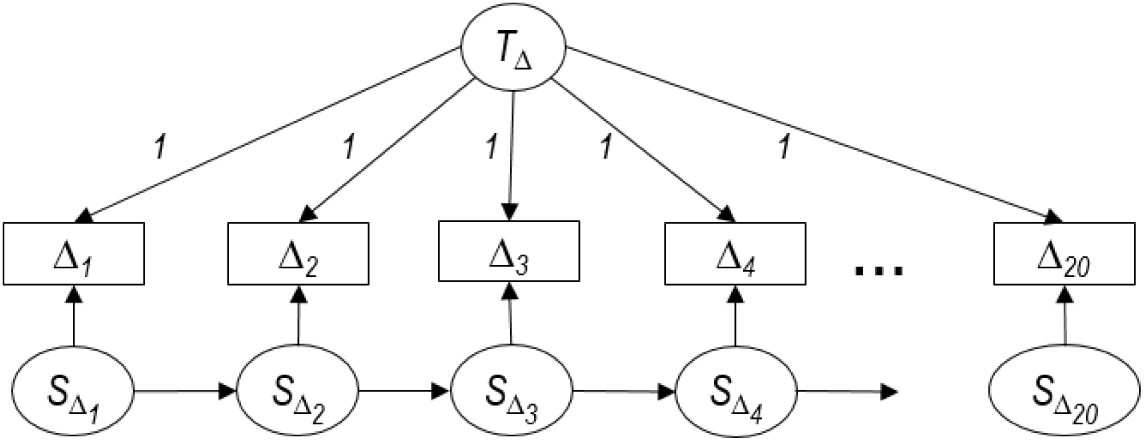
Path diagram of the LST-AR model for one latent variable measured by one indicator (either ΔSNR_a_ or ΔSNR_b_ depending on RQ1) on the 20 measurement occasions of the longitudinal data (*N_2_ = 169*). *T*: latent trait variable; *Δ*: observed indicator; *S*: latent state variable.

#### RQ 3. Can we perform a data-driven, model-based classification of noise and distortion haters using the proposed listening preference measure and longitudinal data?

For the last research question, the longitudinal data frame (*N_2_* = 169) was used. We extended the previous models to mixture LST and mixture LST-AR model. Mixture models aim at detecting latent subgroups of individuals differing in some specified model parameters that drive the distribution of the observed variables (Holtmann et al., 2023). The models were implemented in `Mplus` using the MLR estimator with a varying number of random sets of starting values (between 100/10 and 1000/100 depending on the number of parameters to be estimated). In a first step, we investigated the distribution of latent classes *c* that can be separated based on their trait means, holding all other LST model parameters constant across classes:

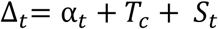

Two-, three-, and four-class solutions were estimated. The evaluation of the optimal solution was based on model fit indices (AIC, BIC, SABIC) and entropy, as well as on the substantive interpretability of the identified classes, as recommended by several authors (e.g., Holtmann et al., 2023; B. Muthén, 2003; Ram & Grimm, 2009).

In a second step, we extended the LST-AR model to a mixture LST-AR model in order to identify latent classes *c* of participants that can be separated based on their listening preferences dynamics, namely with different distributions of LST-AR model parameters:

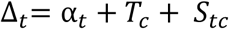

We again estimated two-, three-, and four-class solutions and evaluated them with respect to model fit, entropy and interpretability.

Finally, we explored potential covariates of the latent classes that emerged from the best classification solutions. We used one-way ANOVAs with between-subject factor classes for each of the following variables: average hearing performance (SRT), sound preference and hearing habits (SP-HHQ subscales annoyance/distraction by background noise, importance of sound quality, noise sensitivity), noise sensitivity (WNSS global scale) and personality (NEO-FFI subscales neuroticism, extraversion and conscientiousness scales).

## Results

### Response to RQ 1: Both indicators measure the same rank order and ΔSNR_a_ shows higher consistency

The two latent factors ΔSNR_g_ and ΔSNR_a_ showed a correlation of *r* = .86 in the two-factor model. The model fit of the two-factor solution did not improve over the single-factor model significantly, with 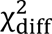(1, *N* = 185) = 1.1, *p* = .293, RMSEA = .024. Moreover, the observations for the two indicators at the same time point showed correlations of magnitude *r* = .40 to *r* = .50. We can therefore conclude that the single-factor model best fits the data, and that the two indicators ΔSNR_g_ and ΔSNR_a_ are measuring the same rank order of individuals.

To decide which of the two indicators (ΔSNR_g_ or ΔSNR_a_) should have been considered for subsequent analysis and as best measurement indicator in a mobile app of listening preferences, we inspected the magnitude of their factor loadings in the single-factor model. As can be seen in figure 6, the loadings for ΔSNR_g_ were on average *λ*_Δg_=.378 and the ΔSNR_a_ loadings were on average *λ*_Δa_=.501. Higher loadings for ΔSNR_a_ onto the factor mean that this indicator had higher overall consistency across repeated measures. In addition, when inspecting the distributions of the single observations (see figure 7), ΔSNR_a_ showed the noise-distortion trade-off more clearly. Indeed, the distribution of the SNR differences showed a median shift of about 4 dB, indicating that participants were successfully “forced” to take a decision in the adaptive trade-off condition created in slider 3. Furthermore, the distribution of accepted clipping ratios was also broader for ΔSNR_a_, leading to a better separation between noise haters preferring to keep their chosen SNR at the cost of high distortion, and distortion haters preferring to reduce distortions at the cost of SNR. In contrast, many observations for ΔSNR_g_ had a negative value, meaning that participants chose a higher SNR level in slider 2 (general trade-off) than in slider 1 (linear gain scenario). These negative values are unexpected, as they indicate that individuals do not adjust slider 2 beyond their preferred SNR position of slider 1, as this would introduce distortion that would impair listening effort. Such cases of negative ΔSNR_a_ might indicate that the level of distortions present in slider 2 at the preferred SNR position of slider 1 was too low to successfully enforce a noise-distortion trade-off.

**Figure 6.**
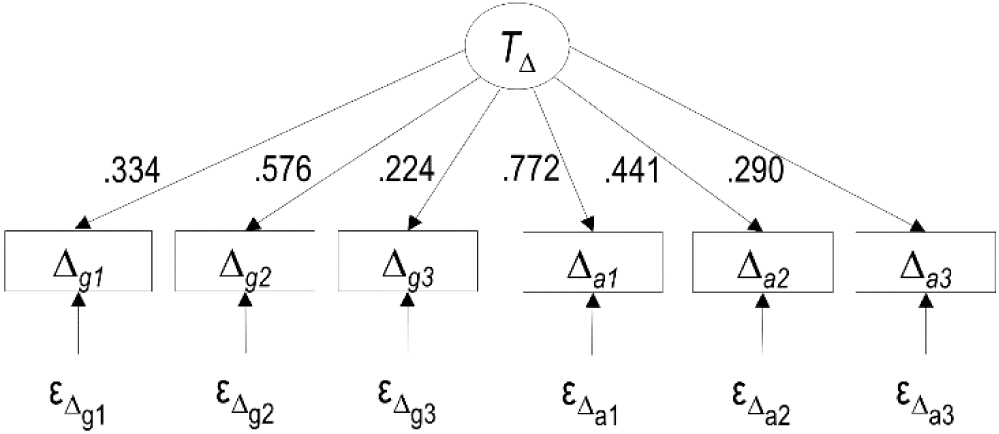
CFA diagram of the single-factor model for the two indicators ΔSNR_g_ (here depicted as Δ_g_) and ΔSNR_a_ (here depicted as Δ_a_) for the three measurement occasions at baseline (*N_1_ = 185*). Factor loadings for each indicator are shown. *T*: latent trait; **λ**: factor loading; *Δ*: observed indicator; ε: measurement error.

**Figure 7.**
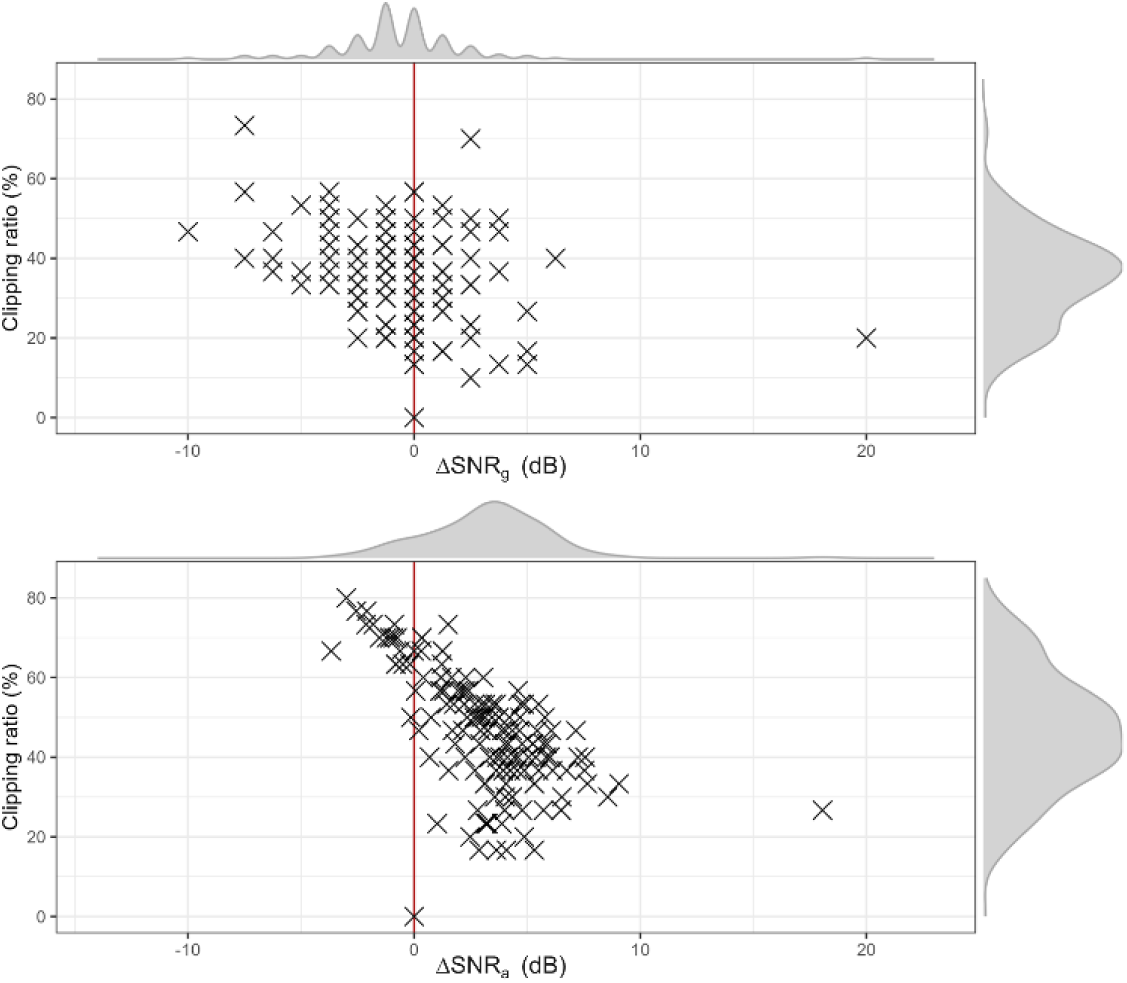
Distribution of observations for ΔSNRg (top) and ΔSNRa (bottom). The X axis shows the ΔSNR in dB measured as the difference between slider 1 - slider 2 and between slider 1 - slider 3. The Y axis shows the corresponding clipping ratio. For each of the *N_1_* = 185 participants the three baseline measures are shown. The marginal density plots show the overall distributions of the individual ΔSNR measures and clipping. SNR: signal-to-noise ratio.

### Response to RQ 2. There is a considerable amount of state-related variance but no temporal dependencies between consecutive observations

First, we implemented the single-trait LST model for ΔSNR_a_ (as chosen in RQ1) to investigate the overall state-related variance. The complete `Mplus` output can be consulted in our Zenodo repository (https://doi.org/10.5281/zenodo.13960717). Table 1 provides an overview of model fit indices, with good model fit usually indicated by RMSEA < .07; CFI > 0.95; TLI > .95; SRMR < .08 and a low χ^2^ relative to the degrees of freedom with *p* > .05 (Hooper et al., 2008). Among all indices, the RMSEA indicated good fit, while the others did not meet the cut-off criteria. The average state-related variance amounted to .787 for the 20 longitudinal measures taken into account (*N_2_* = 169 participants). This indicates that almost 79% of the variance observed in the measured listening preferences is attributable to situational (or state) fluctuations, while about 21% of the variance observed is due to stable trait-like preferences. Variance estimates for the single time points varied between .607 (*p* = < .001) and .895 (*p* = <.001) and can be consulted in the `Mplus` output.

**Table 1.** Overview of average state-related variance and model fit for the LST model and the LST-AR model on the longitudinal data (*N_2_* = 169)

| Model | $S$ | AIC | BIC | SABIC | RMSEA | CFI | TLI | SRMR | $\chi^2$ | df | p-value |
| --- | --- | --- | --- | --- | --- | --- | --- | --- | --- | --- | --- |
| LST | .787 | 15444.777 | 15573.103 | 15443.285 | .065 | .743 | .741 | .102 | 322.432 | 189 | < .001 |
| LST-AR | .778 | 15422.094 | 15609.888 | 15419.911 | .060 | .802 | .779 | .090 | 272.499 | 170 | < .001 |

\*Note: LST: latent state-trait model; LST-AR: autoregressive latent state-trait model; $S$ : average state-related variance; AIC: Akaike information criterion; BIC: Bayesian information criterion; SABIC: sample-size adjusted BIC; RMSEA: root mean square error of approximation; CFI: comparative fit index; TLI: Tucker–Lewis index; SRMR: standardised root mean square residual; $\chi^2$ : Chi-Square; df: degrees of freedom.

Secondly, we extended the LST model for ΔSNR_a_ to a LST-AR model to explore potential time-series dynamics of the observed state-related variance. First-order AR effects indicate the temporal dependency of each observation on the previous one. When taking AR effects into account, the model fit improved. AIC, BIC, SABIC were smaller than in the LST model and the other fit indices were closer to the cut-off values (see table 1). The average state-related variance amounted to .778 (see the `Mplus` output for the single time points estimates). Overall, four out of 19 AR effects were significant: AR effect of time point 2 on 3 (.222, *p* = .004), time point 6 on 7 (.120, *p* = .046), time point 8 on 9 (.243, *p* = < .001), and time point 18 on 19 (.349, *p* = < .001). These four first-order AR effects indicate an effect of the evening measurement on the following morning’s observation. However, these effects had very low magnitude and did not show a consistent pattern, thus no conclusions on systematic linear temporal dependency could be derived. Consequently, we did not estimate second-order AR effects, which refer to the dependence of an observation to the previous-to-last time point (in our case the dependence of a morning measurement on the previous morning and of an evening measurement on the previous evening). We refer to the `Mplus` output for all first-order AR effects estimates (see Zenodo repository).

### Response to RQ 3.1 Data-driven classification suggests the identification of three classes based on the trait means

We extended the analysis to a mixture LST model for ΔSNR_a_. AR effects were not taken into account for classification since only weak effects had been observed (see above). First, we investigated the distribution of latent subgroups or classes *c* that could be separated based only on their trait means *T*. For this, the intercepts *α_t_* were fixed to equality across time and the state-related variance *S_t_* was constrained to equality across classes. Mean differences in latent trait levels were freely estimated and allowed to vary across latent classes. The results of the two-, three-, and four-class solutions can be found in table 2, together with model fit and entropy measures. The two-class solution showed the highest entropy and identified a small class comprising six individuals with trait mean close to zero, which aligns with our definition of noise haters. The remaining participants had been grouped into a second majority class, which may encompass distortion haters of varying degrees. The three-class solution further differentiated this majority class, thereby enhancing the interpretability of the classification output. In addition to the six noise haters (*T*_*c*1_ = 0.643), the three-class solution identified a small class with the highest trait mean (*T*_*c*3_ = 6.055), which may encompass a group of extreme distortion haters. The majority class exhibited here an intermediate mean trait (*T*_*c*2_ = 4.103). The selection of the optimal solution relies on balancing information provided by the metrics (model fit and entropy) and the interpretability of the latent classes. The three-class solution was selected as the optimal solution, as it provided the highest model fit (as indicated by the smallest AIC and SABIC), while maintaining a good level of entropy. It should be noted that model fit indices and entropy are on different scales, which means that even small differences in model fit can motivate the choice of a model over a competing one. Furthermore, discerning three distinct classes (here noise haters, intermediate, and distortion haters) is epistemologically equivalent to identifying two classes where one exhibits varying degrees of the trait. The three-class solution is also consistent with previous findings, wherein laboratory studies similarly identified a comparable distribution of three classes. Figure 8 shows the class-specific distribution of individual ΔSNR_a_ averaged across all longitudinal measures, and illustrates the model-derived classification of individuals into the three distinct classes. It should be noted that the model-derived class-specific trait means do not equal the class-specific arithmetic means of the observed ΔSNR_a_ measures. The latent trait is an estimated variable reflecting an attribute of a person that is not directly measured but inferred from the observed data (Steyer et al., 2015).

**Figure 8.**
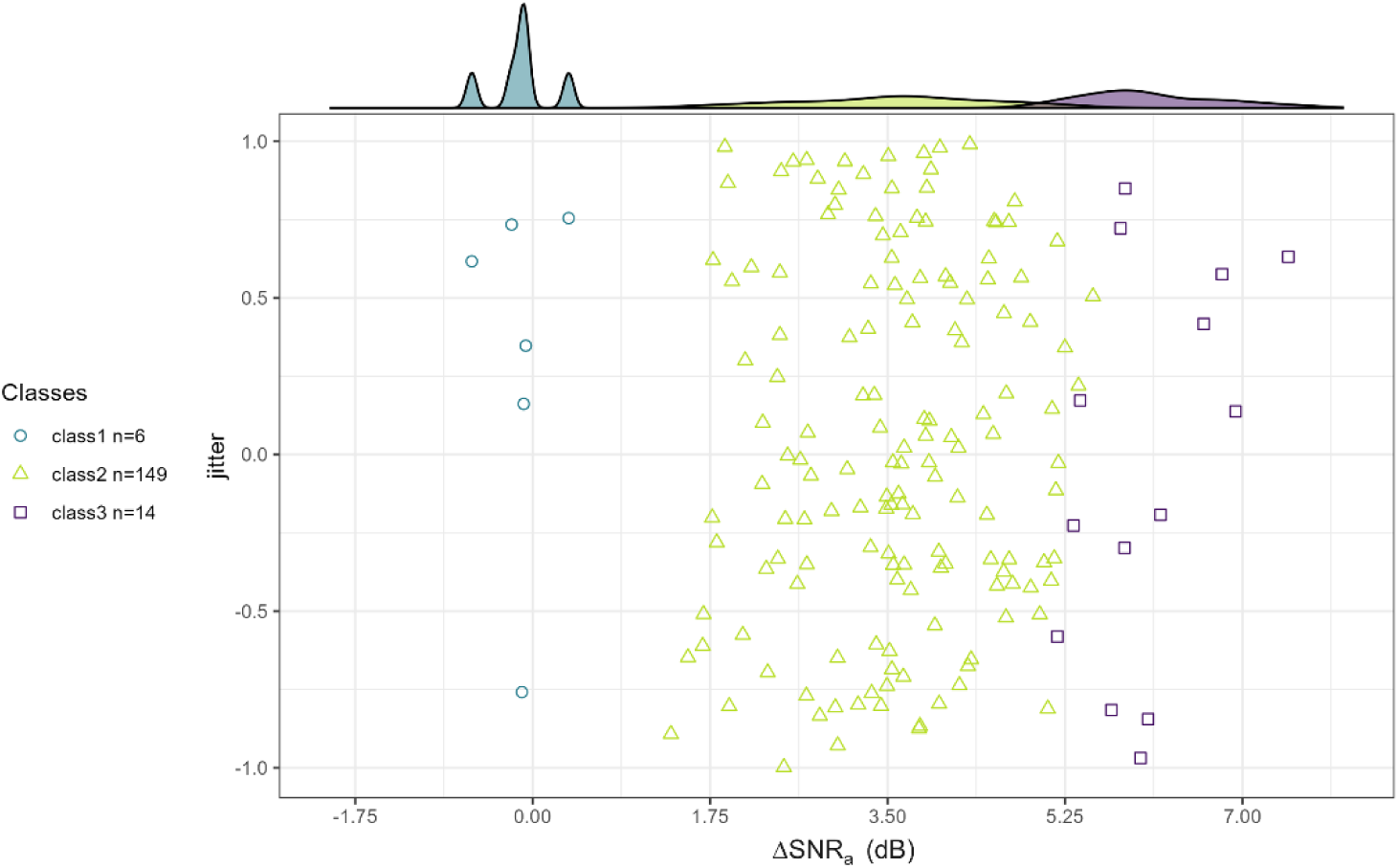
Class-specific distributions of individual mean ΔSNR_a_ averaged across all longitudinal measures for the 3-class solution based on the trait mean (*N_2_* = 169). The three classes are color- and shape-coded. The single measures are jittered along the Y axis for better visualization. The marginal density plot shows the class-specific distributions of the individual mean ΔSNR_a_. SNR: signal-to-noise ratio.

**Table 2.** Overview of model fit, entropy, class sizes and trait means for the mixture LST models with classification based on the trait mean for the longitudinal data set (*N_2_* = 169)

|  | **AIC** | **BIC** | **SABIC** | **Entropy** | **Class sizes** |  | **$T_c$** |
| --- | --- | --- | --- | --- | --- | --- | --- |
| **2 classes** | 15438.856 | 15573.442 | 15437.292 | .946 | class 1 | 6 | 0.759 |
|  |  |  |  |  | class 2 | 163 | 4.354 |
| **3 classes** | 15437.242 | 15578.087 | 15435.604 | .806 | class 1 | 6 | 0.643 |
|  |  |  |  |  | class 2 | 149 | 4.103 |
|  |  |  |  |  | class 3 | 14 | 6.055 |
| **4 classes** | 15439.448 | 15586.553 | 15437.737 | .713 | class 1 | 6 | 0.474 |
|  |  |  |  |  | class 2 | 36 | 3.170 |
|  |  |  |  |  | class 3 | 109 | 4.46 |
|  |  |  |  |  | class 4 | 18 | 6.283 |

\*Note: AIC: Akaike information criterion; BIC: Bayesian information criterion; SABIC: sample-size adjusted BIC; $T_c$ : class-specific trait mean.

### Response to RQ 3.2 Classification improves when accounting for state-related variance

We extended the previous classification (see response to RQ 3.1) to account for the longitudinal dynamics of listening preferences. Due to the small number of individuals classified in the noise hater class (*n_c1_* = 6) and in the distortion hater class (*n_c3_* = 14), the analysis was conducted only on the majority class (*n_c2_* = 149). A mixture LST model was implemented to identify latent subgroups *c* of participants that differed in their state-trait variability. Intercepts *α_t_*, trait mean *T* and state-related variances *S_t_* were freely estimated and allowed to vary across latent classes. Occasion-specific state-related variances were averaged across classes to a single value *S* per class. The results of the two-, three-, and four-class solutions can be found in table 3. The four-class solution reached only a local solution, as the number of parameters was larger than the sample size. Model fit indices and entropy measures favored the three-class solution. This solution identified a majority class with low state-related variance and a comparatively lower trait mean (*T*_*c*1_ = 3.928, *S*_*c*1_ = 0.880). The second class had the highest trait mean and intermediate state-related variance (*T*_*c*2_ = 4.966, *S*_*c*2_ = 0.892). The third class had an intermediate mean and the highest variance (*T*_*c*3_ = 4.089, *S*_*c*3_= 0.943). When accounting for state-related variance, individuals who exhibited similar trait levels could be differentiated based on the degree of variability observed in their listening preferences. Figure 9 shows the class-specific distribution of individual ΔSNR_a_ mean and standard deviation across all longitudinal observations (for *N_c2_* = 149 participants). As with the considerations made for figure 8, it should be noted that the standard deviation of the observed data does not directly correspond to the model-derived state-related variance (which is a standardized latent variable ranging from 0 to 1). In figure 9, standard deviations were employed for the purpose of visualizing the degree of variability observed in the data for each model-derived class.

**Figure 9.**
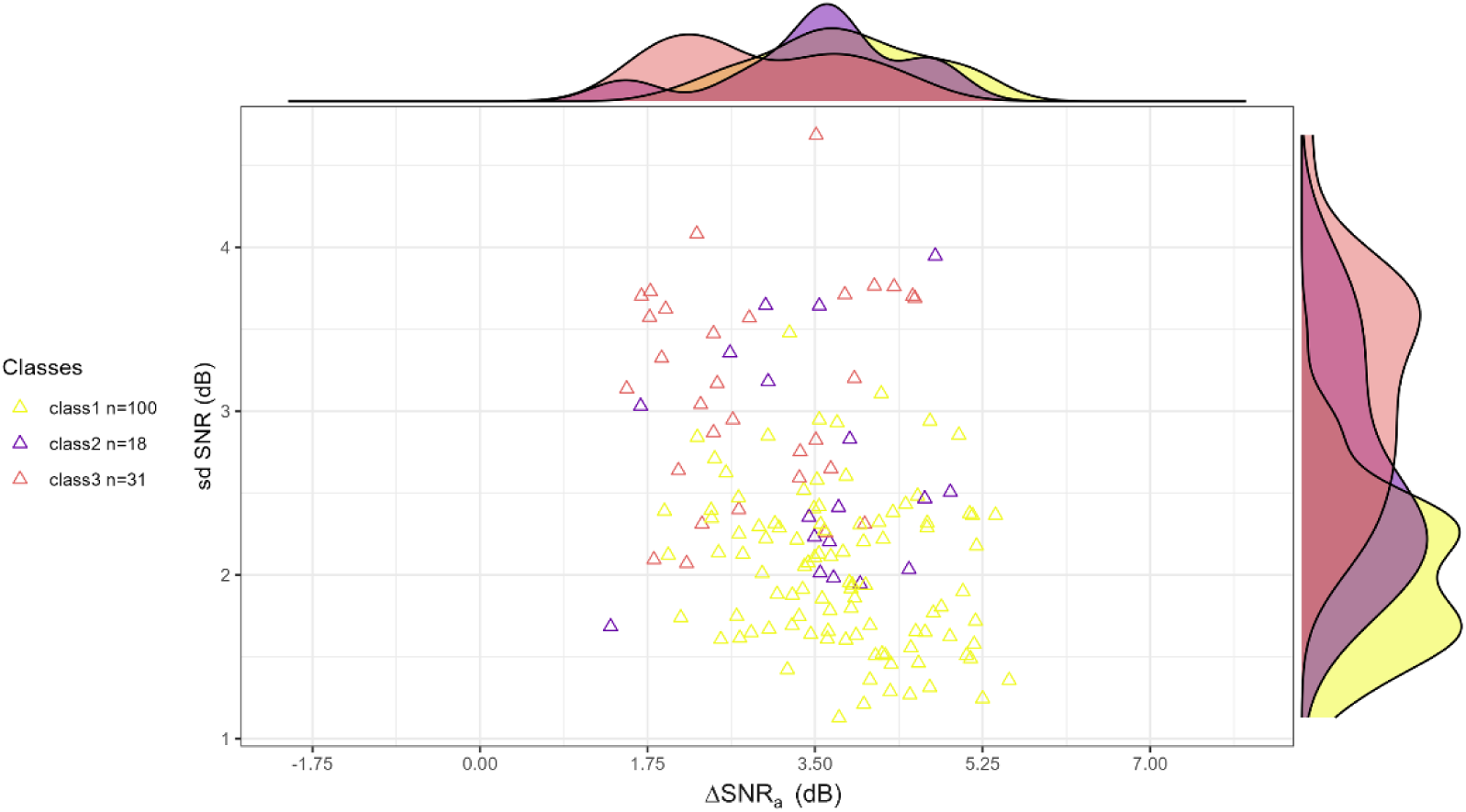
Class-specific distributions of individual mean ΔSNR_a_ (X axis) and standard deviation (Y axis) across all longitudinal measures for the 2-class solution based on LST model parameters (*N_c2_* = 149). Three latent subgroups have been here separated from the majority class identified in the first classification step (RQ 3.1). The classes are color-coded. The marginal density plots show the class-specific distributions of the individual mean ΔSNR_a_ and of the individual standard deviations. SNR: signal-to-noise ratio; *sd*: standard deviation.

**Table 3.** Model fit indices, entropy, class-specific proportions, trait-means and state variances for the mixture LST models with classification based on the LST model parameters for the majority class, identified in RQ 3.1 through classification based on the trait means (*N_cl2_* = 149).

| | AIC | BIC | SABIC | Entropy | Class sizes | | $T_c$ | $S_c$ |
| --- | --- | --- | --- | --- | --- | --- | --- | --- |
| **2 classes** | 13231.513 | 13477.836 | 13218.329 | .849 | class 1 | 110 | 4.015 | 0.885 |
|  |  |  |  |  | class 2 | 39 | 4.303 | 0.948 |
| **3 classes** | 13156.464 | 13525.950 | 13136.689 | .900 | class 1 | 100 | 3.928 | 0.880 |
|  |  |  |  |  | class 2 | 18 | 4.966 | 0.892 |
|  |  |  |  |  | class 3 | 31 | 4.089 | 0.943 |
| **4 classes**<br>*\*$p > N$* | 13122.124 | 13614.771 | 13095.757 | .914 | class 1 | 24 | 3.456 | 0.856 |
|  |  |  |  |  | class 2 | 30 | 3.981 | 0.949 |
|  |  |  |  |  | class 3 | 79 | 4.181 | 0.885 |
|  |  |  |  |  | class 4 | 16 | 4.815 | 0.855 |

\*Note: The four-class model reaches a local solution due to the number of parameters $p$ being larger than the sample size $N$ . In the three-class solution a residual variance of 0 and trait loading of 1 was observed for time point 3. Given the small class size this parameter was not well identified and it was therefore not included when computing the average state variance $S$ of the class. No other anomalies occurred in the model. AIC: Akaike information criterion; BIC: Bayesian information criterion; SABIC: sample-size adjusted BIC; $T_c$ : class-specific trait mean; $S_c$ : class-specific average state variance.

### Response to RQ 3.3 The class with largest state-related variance shows significantly higher neuroticism trait level

We investigated potential covariates for the latent classes identified from the data-driven, model-based classifications (RQ 3.1 and RQ 3.2). An overview of all one-way ANOVAs results is provided in the supplementary material 3. The latent classes identified in response to RQ 3.1 (classification based on trait means) did not significantly differ with respect to hearing performance, sound preference and hearing habits, noise sensitivity and personality. When considering the latent classes identified in response to RQ 3.2 (classification based on trait and state-variance on *N_c2_* = 149), hearing performance and neuroticism only approached significance with *p* < .1. Specifically, the ANOVA results for hearing performance were *F_(2,146)_* = 2.390, *p* = .095 with no significant class differences at the post-hoc pairwise comparisons (TukeyHSD test with 95% CI). Despite the subjective reports of hearing difficulties, 74.1% of all participants had on average good hearing performance (individual mean SRT < −7.1 dB SNR) at the repeated measures of the DTT. 2.7% of participants showed intermediate average performance (individual mean SRT >= −7.1 dB SNR and < −5.1 dB SNR) while the remaining 3.2% had on average poor hearing performance (individual mean SRT >= −5.1 dB SNR) (Buschermöhle et al., 2014; Smits et al., 2006). The ANOVA results for neuroticism were *F_(2,146)_* = 3.630, *p* = .050 and the post-hoc pairwise comparisons (TukeyHSD test with 95% CI) revealed a significant difference between class 2 and 3 (*p* = .043). As can be seen in figure 10, class 3, which had shown the highest state-related variance (*S*_*c*3_= .943), had significantly higher neuroticism values as compared to class 2, which had shown comparatively lower state variance (*S*_*c*3_= .892).

**Figure 10:**
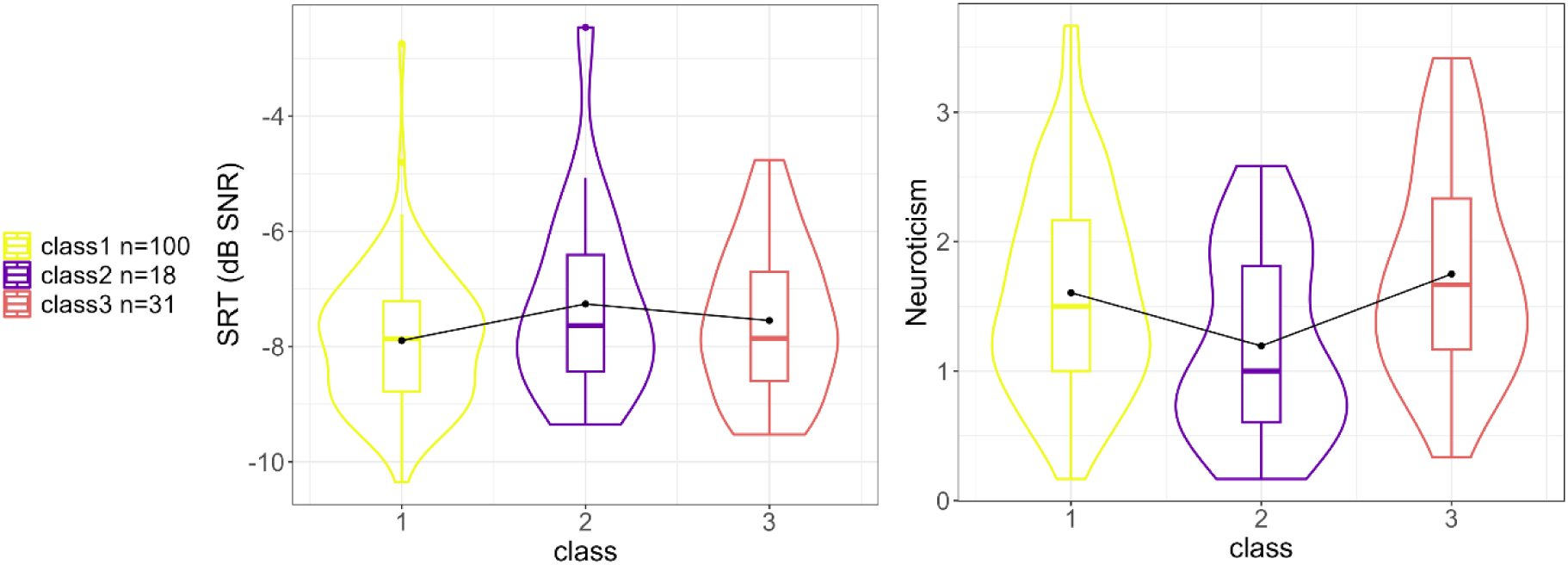
Class-specific distributions of individual SRT mean values (averaged across all longitudinal measures) and individual neuroticism trait levels for the classification based on the LST model parameters for *N_c2_* = 149 (RQ 3.2). The violin plots depict the distribution of all observations within each class, and the box plots represent the median, interquartile range, and potential outliers. SRT: Speech Recognition Threshold, SNR: signal-to-noise ratio.

## Discussion

The present study evaluated the psychometric quality of a newly proposed mobile measure of individual listening preferences along the noise-distortion trade-off and showed that such preferences are not fully stable over time. Through this novel mobile task, we were able to measure individual listening preferences outside the laboratory for 185 older adults with subjective hearing difficulties. Listening preferences were assessed over ten days (mornings and evenings), resulting in a total of 20 measurement time points. The difference between preferred SNR level in a linear gain scenario (slider 1) and in an adaptive noise-distortion trade-off scenario (slider 3) shows good between-person consistency across three consecutive repetitions in a baseline assessment. The adaptive trade-off scenario simulated the general effect of a NR algorithm and the derived measure can help in estimating the individual preference for NR strength. Smaller values (ideally close to zero) on the derived difference measure indicate less tolerance towards noise and a need for stronger NR, characterizing a noise hater (Kubiak et al., 2022; Völker et al., 2018) or NR lover (Neher & Wagener, 2016). Larger values on this difference measure indicate less tolerance towards distortions and characterize a distortion hater (Kubiak et al., 2022; Völker et al., 2018) or NR hater (Neher & Wagener, 2016). This noise-distortion trade-off measure could serve as a preliminary reference point in the fine-tuning stage of a NR algorithm, informing a clinically based selection of a limited pre-set of NR strengths. Our measure relied on a discrete number of signal choices (buttons) for each slider due to limitations in mobile implementation. Nevertheless, the use of a discrete button approach in a mobile task proves sufficient to evaluate the trade-off, with results comparable to those obtained with a continuous slider (Gößwein et al., 2022).

The LST-AR modeling framework was employed to investigate the collected set of longitudinal data, with the aim of evaluating trait consistency and state specificity of individual preferences for noise vs. distortion. These preferences, which had been so far regarded as a stable individual trait (Kubiak et al., 2022; Neher & Wagener, 2016; Reinten et al., 2023; Völker et al., 2018), show considerable day-to-day fluctuations within-person. A large amount of the observed variance is attributable to situational states and/or person-situation interaction effects. There is however no temporal dependency between consecutive time points. It follows that the HA fine-tuning process of NR algorithms should account not only for inter-individual differences in listening preferences at supra-threshold levels (Jepsen & Dau, 2011; Kubiak et al., 2022), but also for intra-individual differences in similar listening scenarios. Repeated assessments of listening preferences in everyday life with an easy and short mobile task should be considered for successful fine-tuning of HAs, particularly for those listeners where greater instability is anticipated.

Information on mean trait levels and state-related variances was used to improve on the established classification of individuals into noise haters/NR lovers vs. distortion haters/NR haters (Houben et al., 2023; Kubiak et al., 2022; Neher & Wagener, 2016; Völker et al., 2018). As a first step, we sought to identify latent subgroups of individuals who differed solely in their habitual listening preferences (latent trait means) in order to replicate previous studies that did not consider longitudinal assessments. Three distinct classes emerged. Two comparatively small classes are distinguished by low (noise haters) and high (distortion haters) trait levels, respectively. The majority of individuals are classified into an intermediate class. It could be argued that the most extreme classes, characterized by very low sample sizes, might be influenced by outliers. However, we mitigated this risk by excluding individuals with invariant or inattentive responses (for example, participants who consistently selected the leftmost slider position at −7 dB SNR). Moreover, this distribution of class membership is consistent with previous laboratory studies (Gößwein et al., 2022; Kubiak et al., 2022; Reinten et al., 2023) in which only a limited number of participants exhibited a clear preference for noise or distortion. These findings may indicate that individual preferences along the noise-distortion trade-off, which underlies the preferences for NR strength, do not conform to a binary classification, but rather lie along a continuum, which we also found with our discrete slider measure.

To account for the substantial intra-individual variability observed, we explored a novel classification based on state-related variance. The majority and intermediate class identified before was thus further separated into three subgroups, each differing in their listening preferences dynamics (with different LST model parameter distributions). When no differentiation could be made based on habitual preferences (latent traits), extending the classification to consider the degree of variability in listening preferences proves informative. Knowledge on the individual stability vs. instability of listening preferences can facilitate further personalization of the fine-tuning process of NR algorithms. Moreover, we found that individuals with more unstable listening preferences show higher levels of neuroticism trait. This fits to literature showing neuroticism to be typically linked to instability (Robinson & Tamir, 2005). Indeed, individuals higher in neuroticism exhibit increased variability in daily emotions (Mader et al., 2023), behaviour (Geukes et al., 2017) and basic cognitive processes involved in linking stimulus to response (i.e., reaction times (Robinson & Tamir, 2005)). Moreover, neuroticism has been correlated with more mind-wandering during cognitive tasks, lower working memory capacity, and poorer attention control (Robison et al., 2017).

In the present study, hearing performance does not emerge as a significant covariate. It is important to note, however, that the majority of participants demonstrate good average hearing performance, despite reporting subjective hearing difficulties in daily life. A better understanding of the relationship between hearing performance and listening preferences (and their variability) may be achieved through future studies that include individuals with varying degrees of hearing loss. Previous studies have found an effect of hearing loss on preferred NR strength, showing that hearing impaired listeners prefer greater NR (Houben et al., 2023; Neher & Wagener, 2016) and are less sensitive to distortions (Brons, Dreschler, et al., 2014). Moreover, individuals with greater hearing impairment exhibit larger day-to-day variability of hearing performance in stable listening environments (Kuhlmann et al., 2023). Therefore, it can be hypothesized that individuals with worse hearing performance may prefer stronger NR (noise haters) and show a higher degree of variability in their listening preferences.

### Outlook and limitations

In a subsequent stage of this study, we plan to investigate the determinants of the observed large state-related variance of listening preferences along the noise-distortion trade-off. Other subjective states such as momentary mood or stress might have an influence on the intra-individual variability of listening preferences. We plan to examine cross-lagged effects between state variance in listening preferences and variability in daily mood and hearing performance. Listening preferences may vary depending on the nature of the task and future studies might consider more naturalistic target speech signals. It is possible that listeners who accept distortions applied to a short sentence (as in the present study) may not exhibit similar listening preferences when the distortion is applied to a longer signal, such as an audiobook excerpt or a conversation between different speakers (Walravens et al., 2020). To enhance the ecological validity of the signals, it would be beneficial to consider different types of distortions that better simulate the signal distortion typically created by NR algorithms, such as “musical” noise distortion (Gößwein et al., 2023). Future studies should also assess listening preferences in different populations: children, younger and older adults with different types and degrees of hearing loss, as well as HAs users. Another limitation of the present mobile study is the lack of control over the surrounding environment and measurement conditions. To increase control over the procedures, it would be advisable to install accompanying apps that record the ambient noise and check for the usage of headphones, which should ideally be calibrated. Future feasibility studies are needed to improve the user-interface and the implementation of the noise-distortion preference task in a mobile app. The use of a continuous slider (Gößwein et al., 2022) has the potential to enhance precision in the measurements, with the requisite degree of precision warranting further investigation.

## Conclusion

The present study introduced a novel mobile measure of listening preferences that allows for repeated assessment in an ecological and momentary manner. We evaluated individual listening preferences over several days, shedding light on the large within-person variability of preferences that had been so far considered as a subjective trait stable over time. We highlight how both trait levels and state-variance components should be considered in the classification of listeners, thus improving over the classical view of noise haters and distortion haters and considering the individual’s stability or instability of listening preferences. Gaining knowledge on how individual listening preferences vary between and within individuals can enhance the efficacy of HA self-adjustment programs, particularly in the presence of environmental noise, for individualized HA fitting.

## Data Availability

The data is available upon motivated request to the corresponding authors. Analyses scripts are available in the Zenodo repository.

https://doi.org/10.5281/zenodo.13960717

## Acronyms

AIC: Akaike Information Criterion
ANL: Acceptable Noise Level
AR: Autoregressive
BIC: Bayesian Information Criterion
CFA: Confirmatory Factor Analysis
CFI: Comparative Fit Index
EMA: Ecological Momentary Assessment
HA: Hearing Aid
LST: Latent State-Trait
LST-AR: Latent State-Trait Autoregressive
MLR: Maximum Likelihood Robust
NR: Noise Reduction
PSM: Perceptual Similarity Measure
PTA: Pure Tone Average
RMSEA: Root-Mean-Square Error of Approximation
SABIC: Sample-Size Adjusted Bayesian Information Criterion
SNR: Signal-to-Noise Ratio
SRMR: Standardised Root Mean square Residual
SRT: Speech Recognition Threshold
TLI: Tucker-Lewis Index

## Acknowledgements

We thank all the participants who took part in this study and the Universities that helped us with recruitment: University of Oldenburg, Gasthörstudium; Goethe-Universität Frankfurt am Main, Universität des 3. Lebensalters; Freie Universität Berlin, GasthörerCard Programm; University of Köln, Gasthörer-und Seniorenstudium; University of Kassel, Gasthörendenprogramm; University of Bielefeld, Studieren ab 50. Preliminary results of this research were presented on symposia of the “Hearing4All” Cluster of Excellence, on the Virtual Conference of Computational Audiology (VCCA) 2023 and 2024, on the Hearing Across the Lifespan (HeAL) Conference 2024 and on the German Society for Psychology (DGPs) Congress 2024.

## Statements and Declarations

### Ethical considerations

The study protocol was approved by the Research Ethics Committee of the Carl von Ossietzky Universität of Oldenburg (08.09.2021, EK/2020/020-01).

### Consent to participate

Written informed consent was provided before the beginning of the study. The informed consent was approved by the research Ethics Committee.

### Consent for publication

Not applicable.

### Data availability

Analyses scripts are available at https://doi.org/10.5281/zenodo.13960717. The data is available upon motivated request to the corresponding authors.

### Contributorship

GA, AH and MB conceptualized the study and were involved in protocol development, study design and data analysis. JAG was involved in the design of the measurement task and data analysis. BK contributed to study conceptualization and obtained funding. GA was responsible for participants’ recruitment, data collection and wrote the first draft of the manuscript. All authors reviewed and edited the manuscript and approved the final version of the manuscript.

