## Supplementary figures and images for "Towards an extended classification of noise-distortion preferences by modeling longitudinal dynamics of listening choices"

### Supplementary material 1

#### Model-optimized clipping values

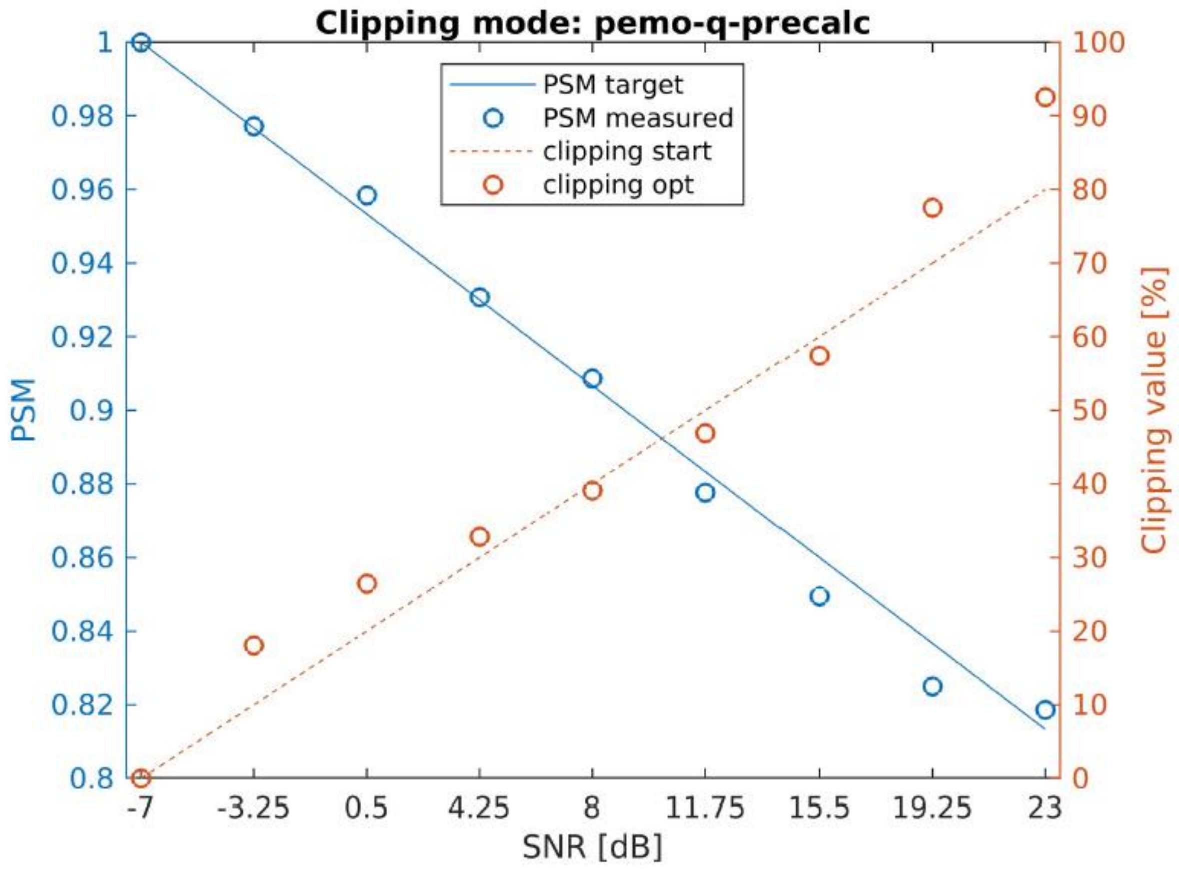

### Supplementary material 2

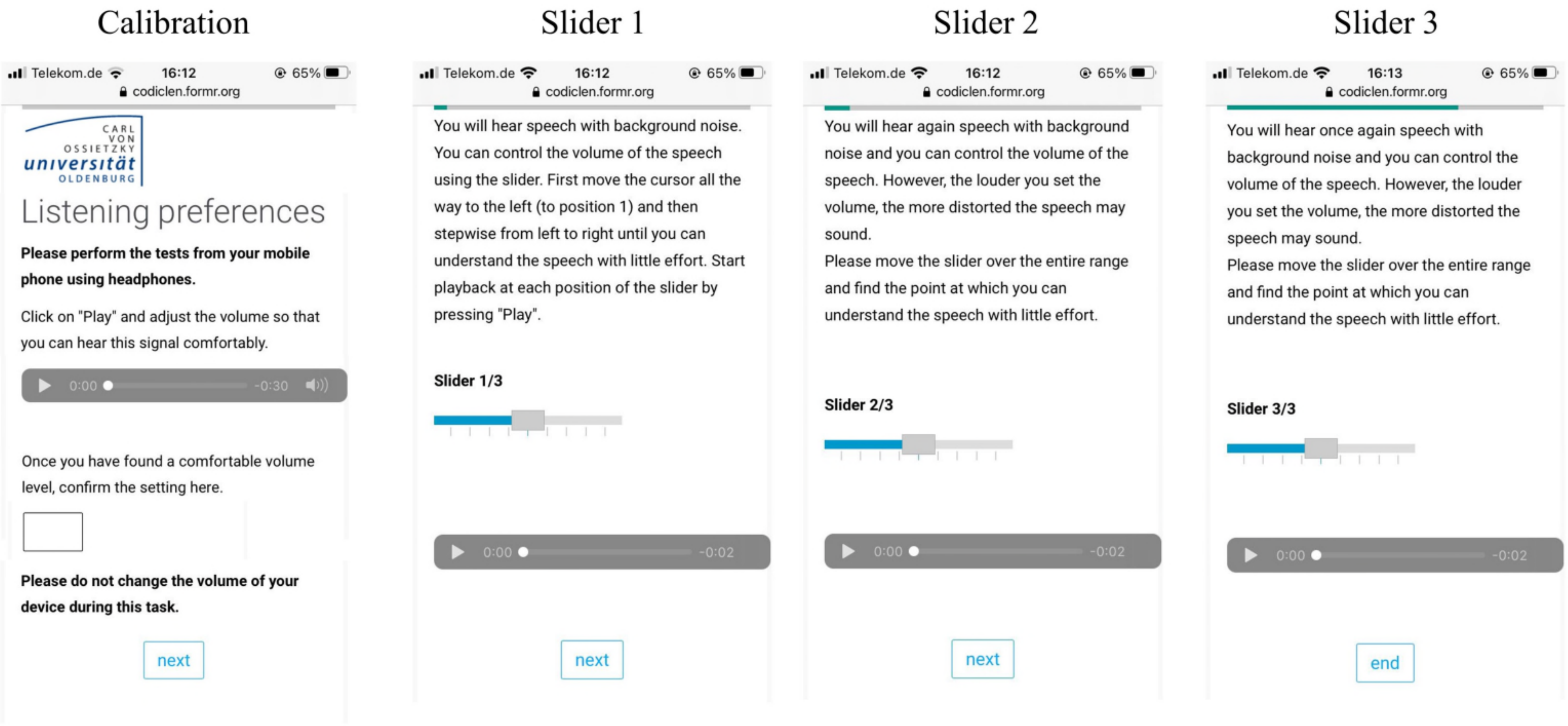
