## Supplementary material 3 for "Towards an extended classification of noise-distortion preferences by modeling longitudinal dynamics of listening choices"

### ANOVA results

##### RQ 3.1) Classification based on the trait means

| Covariate | Df | Sum Sq | Mean Sq | F value | Pr(>F) |
|---|---:|---:|---:|---:|---:|
| **SRT** | (2, 166) | 2.79 | 1.395 | 0.807 | 0.448 |
| **WNSS** | (2, 166) | 150.00 | 75.150 | 0.245 | 0.783 |
| **SP-HHQ f1** | (2, 166) | 53.00 | 26.690 | 0.829 | 0.438 |
| **SP-HHQ f2** | (2, 166) | 1.60 | 0.798 | 0.167 | 0.846 |
| **SP-HHQ f3** | (2, 166) | 4.30 | 2.157 | 0.206 | 0.814 |
| **Neuroticism** | (2, 166) | 0.45 | 0.227 | 0.371 | 0.690 |
| **Conscientiousness** | (2, 166) | 0.59 | 0.296 | 1.246 | 0.290 |
| **Extraversion** | (2, 166) | 0.66 | 0.329 | 1.246 | 0.290 |

##### RQ 3.2) Classification based on the trait means and state variances for the majority class previously identified in RQ 3.1

| Covariate | Df | Sum Sq | Mean Sq | F value | Pr(>F) |
|---|---:|---:|---:|---:|---:|
| **SRT** | (2, 146) | 7.68 | 3.841 | 2.39 | 0.095 |
| **WNSS** | (2, 146) | 152 | 76.18 | 0.241 | 0.786 |
| **SP-HHQ f1** | (2, 146) | 35 | 17.50 | 0.539 | 0.585 |
| **SP-HHQ f2** | (2, 146) | 2.8 | 1.414 | 0.281 | 0.756 |
| **SP-HHQ f3** | (2, 146) | 2.9 | 1.439 | 0.14 | 0.869 |
| **Neuroticism** | (2, 146) | 3.63 | 1.8148 | 3.044 | 0.050 |
| **Conscientiousness** | (2, 146) | 1.04 | 0.522 | 2.255 | 0.108 |
| **Extraversion** | (2, 146) | 0.54 | 0.271 | 1.003 | 0.369 |
